# BrainSignsNET: Deep Learning-Based 3D Anatomical Landmark Detection in Human Brain Imaging

**DOI:** 10.1101/2025.07.31.25332457

**Authors:** Siavash Shirzadeh Barough, Catalina Ventura, Murat Bilgel, Marilyn S. Albert, Michael I. Miller, Abhay Moghekar

## Abstract

Accurate detection of anatomical landmarks in brain Magnetic Resonance Imaging (MRI) scans is essential for reliable spatial normalization, image alignment, and quantitative neuroimaging analyses. In this study, we introduce BrainSignsNET, a deep learning framework designed for robust three-dimensional (3D) landmark detection. Our approach leverages a multi-task 3D convolutional neural network that integrates an attention decoder branch with a multi-class decoder branch to generate precise 3D heatmaps, from which landmark coordinates are extracted. The model was trained and internally validated on T1-weighted Magnetization-Prepared Rapid Gradient-Echo (MPRAGE) scans from the Alzheimer’s Disease Neuroimaging Initiative (ADNI), the Baltimore Longitudinal Study of Aging (BLSA), and the Biomarkers of Cognitive Decline in Adults at Risk for AD (BIOCARD) datasets and externally validated on a clinical dataset from the Johns Hopkins Hydrocephalus Clinic. The study encompassed 14,472 scans from 6,299 participants, representing a diverse demographic profile with a significant proportion of older adult participants, particularly those over 70 years of age. Extensive preprocessing and data augmentation strategies, including traditional MRI corrections and tailored 3D transformations, ensured data consistency and improved model generalizability. Performance metrics demonstrated that on internal validation BrainSignsNET achieved an overall mean Euclidean distance of 2.32 ± 0.41 mm and 94.8% of landmarks localized within their anatomically defined 3D volumes in the external validation dataset. This improvement in accurate anatomical landmark detection on brain MRI scans should benefit many imaging tasks, including registration, alignment, and quantitative analyses.

## Introduction

Accurate localization of anatomical landmarks in human brain scans is essential for numerous clinical and research applications. These landmarks play a crucial role in tasks such as spatial normalization, image alignment, and morphometric analysis, thereby facilitating precise measurements and evaluations in neuroimaging. For instance, in the radiologic assessment of Normal Pressure Hydrocephalus (NPH), key measurements like the Evans Index (EI)(1, 2) and Callosal Angle (CA)(3, 4) require MRI scans to be aligned using Anterior Commissure (AC) and Posterior Commissure (PC). Furthermore, the extraction of specific brain slices, for example, the coronal slice at the level of the PC for CA measurement or delineation of the anterior horn of the lateral ventricles separated by the intraventricular foramen (Monroe) for EI computation, underscores the clinical relevance of precise landmark detection.

In addition to these applications, other central and peripheral landmarks (e.g., anterior and posterior corpus callosum, basal ganglia nuclei, pituitary, optic chiasm, medial temporal lobe structures, and brainstem regions) provide invaluable information for assessing midline shifts, asymmetry between hemispheres, and overall brain morphology.(5–7) These landmarks not only aid in structural alignment and registration but also serve as critical references in lesion localization. For example, detecting the genu of the internal capsule—which demarcates descending neural fibers associated with limb movement—can enhance lesion localization in stroke evaluation, thereby contributing to more clinically actionable insights.(8)

Recent advances in deep learning, particularly the development of three-dimensional (3D) convolutional neural networks (CNNs)(9), have revolutionized medical image analysis.(10) These models have demonstrated the capability to capture intricate spatial relationships within volumetric data, significantly reducing the time and effort required for manual interpretation while improving diagnostic accuracy. In this context, our study introduces a novel 3D deep learning framework that integrates heatmap-based landmark detection for the accurate localization of critical anatomical landmarks in the human brain.

Previous studies have primarily concentrated on anatomical segmentation using both atlas-based and deep learning techniques,(11–13) with limited emphasis on the localization of specific non-volumetric anatomical landmarks. Existing landmark localization methods often operate only on specific types of scans and are vulnerable to noise, variations in image quality, and the presence of abnormalities.(14–19) These limitations significantly restrict their clinical and research utility. To overcome these challenges, our method integrates advanced 3D CNN architectures with heatmap-based localization techniques, providing robust and accurate landmark detection. This framework not only improves accuracy and computational efficiency but also serves as a pretrained model that is trained on large-scale datasets and can subsequently be fine-tuned for diverse applications, including the detection of lesions, abnormalities, and other clinically relevant features in 3D imaging modalities such as CT, PET, and 3D mammography.

In summary, this work presents a state-of-the-art deep learning approach for the detection of neuroanatomical landmarks in the human brain. Our framework offers a robust and precise tool for neuroanatomical analysis, with significant implications for clinical diagnostics and research.

## Methods

### Data Acquisition

T1-weighted MRI data for pretraining, training, and internal validation were drawn from ADNI(20), BLSA(21), and BIOCARD. BLSA scans comprised: at 1.5 T, SPGR acquired on three GE Signa systems(0.94 × 0.94 × 1.5 mm voxels; flip angle 45°; TE 5 ms; TR 35 ms; TI 0 ms) plus one Philips Intera 1.5 T SPGR system with identical parameters, and at 3 T, sagittal MPRAGE acquired on three Philips Achieva systems (1 × 1 × 1.2 mm voxels; flip angle 8°; TE 3.2 ms; TR 6.5–6.8 ms; TI 0 ms). BIOCARD scans included a single 1.5 T GE Genesis Signa SPGR (axial; 1 × 1 × 2 mm voxels; flip angle 20°; TE 2 ms; TR 24 ms; TI 0 ms) and a single 3 T Philips Achieva MPRAGE (1 × 1 × 1.2 mm; flip angle 8°; TE 3.1 ms; TR 6.75 ms; TI 0 ms). For external validation, 200 T1-weighted scans were acquired at the Johns Hopkins Hydrocephalus Clinic on a Siemens Verio 3 T (0.78 × 0.78 × 0.80 mm voxels; flip angle 9°; TE 3.13 ms; TR 1800 ms; TI 900 ms). All images were deidentified by stripping private metadata and converted to NIfTI format using the dcm2niix Python package.

### Annotation

MRI scans from ADNI, BLSA, BIOCARD, and the Johns Hopkins Hydrocephalus Clinic underwent region-of-interest segmentation utilizing the FreeSurfer recon-all pipeline, augmented with specialized flags and functions to accurately segment large ventricles, brainstem regions, and hypothalamic subunits. This approach yielded detailed (but noisy) three-dimensional volumetric representations of the targeted anatomical structures, including the posterior and anterior corpus callosum, right and left putamen, left and right caudate, left and right thalamus, left pallidum, left hippocampus, left amygdala, right pallidum, right hippocampus, right amygdala, optic chiasm, intraventricular foramen, midbrain, pons, medulla, posterior commissure, anterior commissure, and the left and right internal capsule genu. Subsequently, image processing techniques were applied to identify anatomical landmarks within each scan. For volumetric structures such as the thalamus, brainstem, and basal ganglia nuclei, we developed a search algorithm to determine the optimal location for inscribing the largest possible sphere within the given structure (Figure 1a).

**Figure 1.**
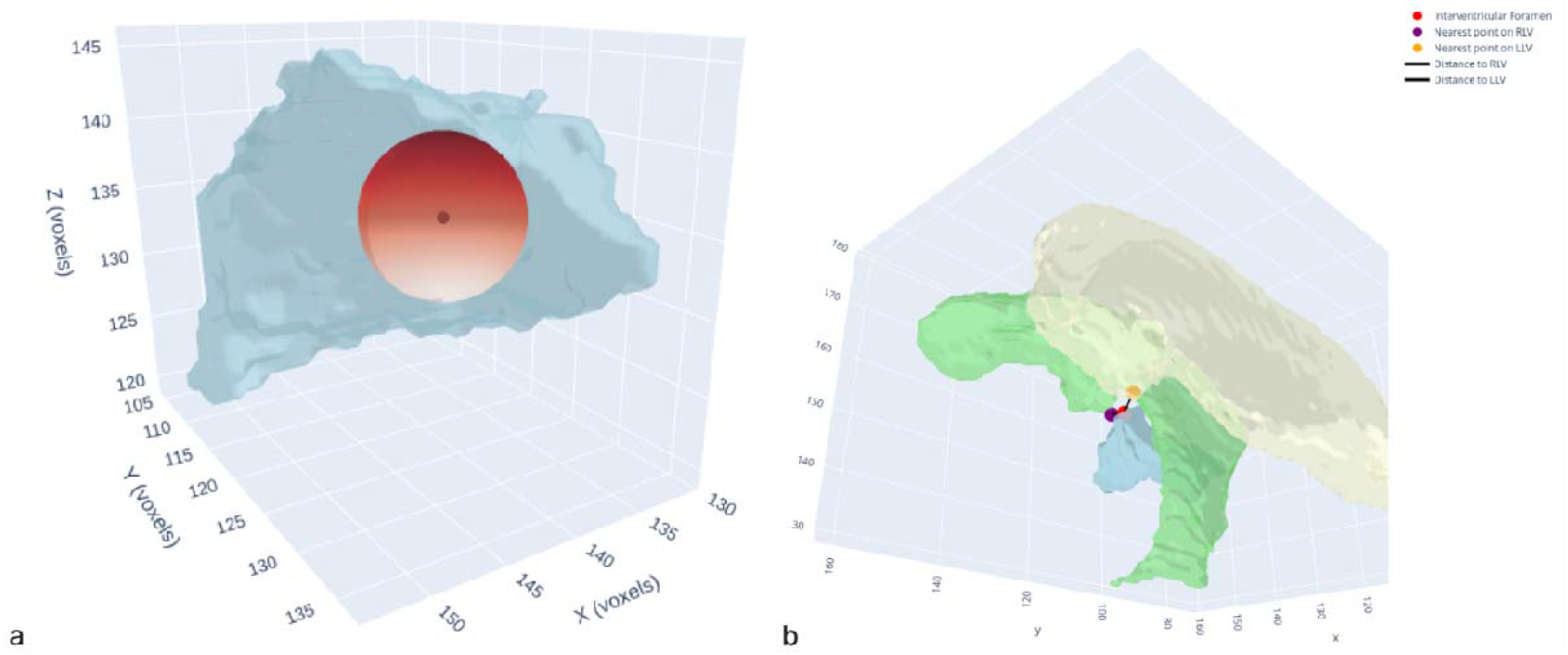
3D illustration of image processing methods used for extracting anatomical landmarks from volumetric segmentation masks. (a) Landmark extraction from volumetric anatomical structures (e.g., the thalamus) by determining the largest possible inscribed sphere within the structure. (b) Landmark extraction by identifying coordinates representing the closest proximity between two adjacent anatomical structures, exemplified here by the intraventricular foramen as the nearest points between the lateral and third ventricles.

**Figure 2.**
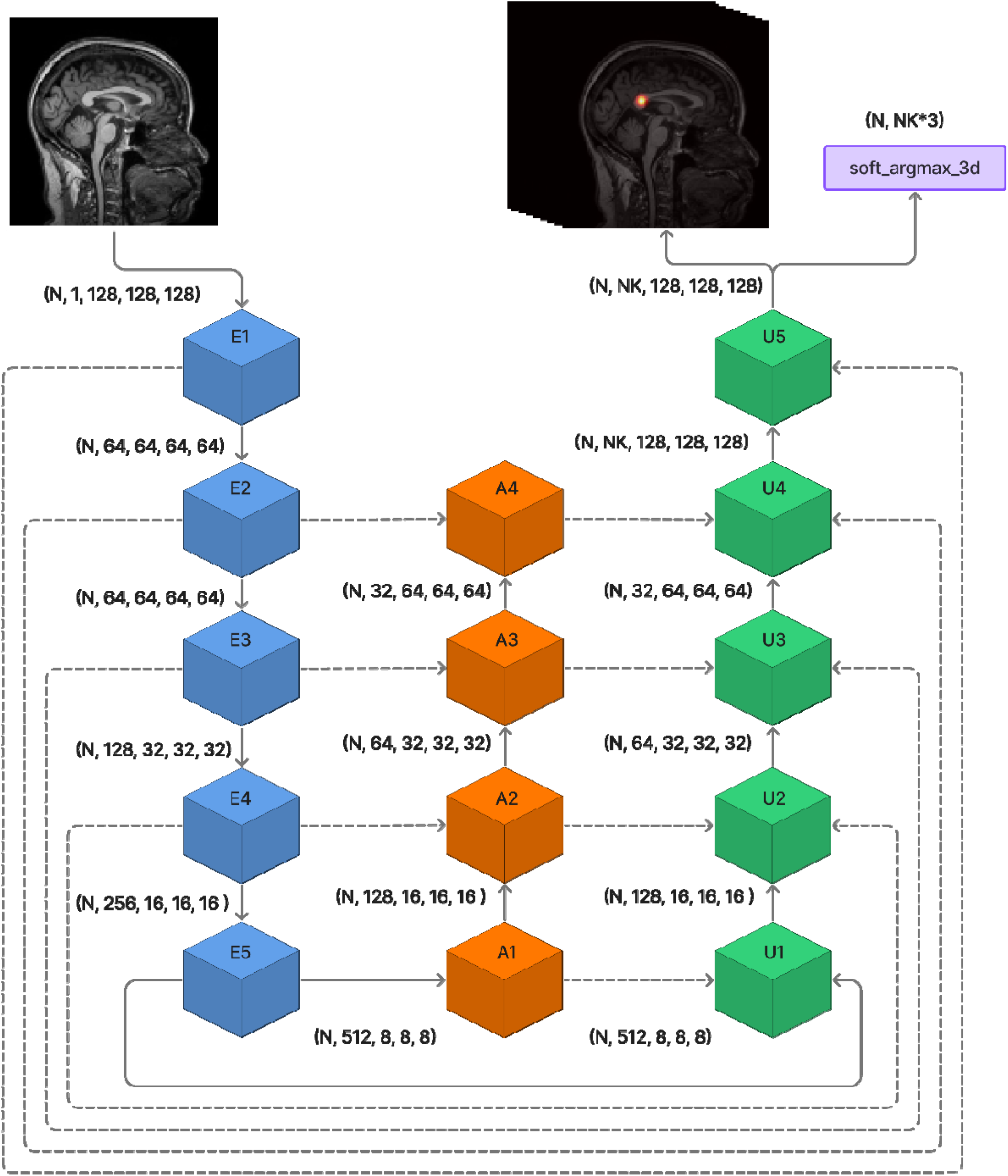
Overview of the proposed model architecture. Blue cubes represent encoder blocks, orange cubes indicate attention blocks, and green cubes depict multiclass heatmap decoder blocks. Solid lines illustrate direct connections, while dashed lines denote skip connections facilitating information transfer between encoder and decoder blocks. Interpolate layers are indicated by R in the figures and are used to resize the multidimensional tensors to be suitable for use as skip connections. N: Batch Size, NK: Number of Landmark.

For non-volumetric anatomical landmarks, spatial relationships between structures were leveraged to determine landmark positions. For instance, as demonstrated in Figure 1b, the intraventricular foramen was identified as the point on the third ventricle that minimizes the distance to both lateral ventricles. Similarly, the genu of the internal capsule was delineated by locating the region within the white matter map that exhibits approximately equal distances from the thalamus, caudate nucleus, and ventral diencephalon. Distances were calculated using the 3D Euclidean distance formula. Finally, all annotations for the BLSA, BIOCARD, and Johns Hopkins Hydrocephalus Clinic datasets were manually reviewed and corrected as needed.

### Preprocessing and Augmentation

To ensure spatial consistency, all scans were first reoriented to the Right–Anterior–Superior (RAS) coordinate frame. We then resampled each volume by computing axis-specific scale factors from the native voxel and applying trilinear interpolation: each dimension’s voxel count is adjusted in direct proportion to its original spacing so that the physical size of the brain remains unchanged and no geometric distortion is introduced. Finally, all image intensities were normalized (zero mean, unit variance) to standardize contrast across scans. To enhance the model’s robustness and generalizability—and to reduce its reliance on any single preprocessing algorithm—we incorporated traditional MRI preprocessing techniques as data augmentation during training.

These techniques included bias field correction, denoising, and skull stripping. In addition, we implemented three-dimensional image augmentation functions. These functions comprised rotations along multiple axes, random cropping, random zooming, as well as shifts in intensity, brightness, and contrast. Random Gaussian noise and elastic deformations were also applied, all while preserving the spatial integrity of the annotated anatomical landmarks.

### Model Architecture

We applied a multi-task 3D landmark detection network that integrates a 3D Convolutional Neural Network backbone with two complementary branches: an attention branch and a multi-class decoder branch generating individual landmark heatmaps. The network predicts 3D heatmaps, from which landmark coordinates are extracted.

The encoder employs a pretrained 3D ResNet-18 model; The Intermediate feature maps from the encoder’s stem and subsequent blocks are preserved. These encoder features are transformed via 1×1 convolutions to serve as skip connections, facilitating multi-scale feature fusion in both decoder branches.

In the attention branch, spatial resolution is progressively restored using four upsampling stages, each comprising a 3D transposed convolution followed by convolutional layers with batch normalization and ReLU activations. Skip connections from encoder blocks are integrated at corresponding resolutions to enrich spatial details.

The multi-class decoder branch similarly reconstructs spatial resolution through four upsampling stages. Each stage combines transposed convolutions, convolutional blocks, and skip connections from both the encoder and the corresponding blocks of the attention branch. Specifically, attention branch outputs at matched resolutions are combined with encoder skip features to enhance localization accuracy. A final convolutional head generates full-resolution 3D heatmaps individually for each landmark.

The soft-argmax-3d operation(22) we employ first flattens each K-channel heatmap into a 1D array and applies a numerically stable, log-sum-exp-backed softmax, yielding a smooth probability distribution over all voxels. By computing an expectation of the coordinate indices, we obtain continuous, differentiable landmark predictions that support back-propagation:

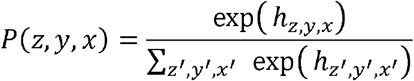

The expected 3D coordinates for each landmark are computed as the weighted average of grid indices:

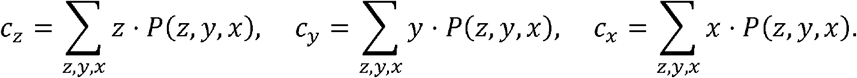

### Training

The training process consisted of several key stages. Initially, all images underwent a standardized preprocessing pipeline. Simultaneously, coordinate data were adjusted to account for the aspect ratio and dimensions of each scan, leveraging anatomical information derived from FreeSurfer-generated 3D ROI volumes or manually annotated landmarks.

Subsequently, data augmentation was applied randomly to the paired scan/coordinate data. This augmentation strategy ensured that the network was exposed to a variety of transformed inputs, enhancing its robustness and generalizability.

Following augmentation, three-dimensional heatmaps were generated for each landmark by applying an isotropic Gaussian blur with *σ* = 2 *pixels* to the binary mask (consisting of a single ON voxel) for each landmark.

To enhance landmark-localization precision, we employed a composite loss framework that jointly optimizes multiclass heatmap reconstruction and coordinate regression. In each training epoch, we compute two Mean Square Error (MSE) losses: one measuring the discrepancy between predicted and ground-truth heatmaps, and the other measuring the error between predicted and true landmark coordinates. We balance these terms with weighting factors—assigning higher weight to the heatmap loss to establish accurate global spatial context as the main training loss. Near convergence, both losses are scaled to comparable magnitudes so that our checkpointing criterion selects models excelling both in heatmap and coordinate accuracy. This strategy sharpens heatmap peaks around true landmark centers and ensures that the final model checkpoint delivers optimal landmark-detection performance rather than focusing on heatmap quality alone. The overall loss was defined as:

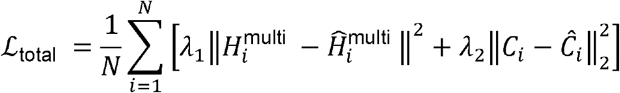

Where 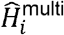 and 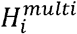 represent the ground-truth and predicted multiclass heatmaps, respectively, obtained by flattening volumetric matrices of size 128 × 128 × 128 into one-dimensional vectors, *Ĉ*_*i*_ and *C*_*i*_ denote ground-truth and predicted landmark coordinates, and N indicates the total number of landmarks. The weighting factors λ_l_, and *λ*_2_ control the relative contribution of each loss component during model training. Weighting factors were selected after a trial training session to make sure that both heatmap and coordinate losses have similar convergence magnitudes.

Given that the selected cohorts included multiple MRI scans per participants, the dataset was partitioned by participant IDs (80% for training and 20% for internal validation) to prevent data leakage and overfitting (for both internal and external validation datasets only one scan per participant was used). Training progress was monitored using per-epoch metrics such as the mean Euclidean distance between predicted and true landmarks.

We employed the AdamW optimizer with weight decay alongside a StepLR scheduler to systematically reduce the learning rate at predefined intervals. Throughout training, we tracked the combined validation loss and saved model checkpoints whenever it improved. We defined convergence as the point at which the validation loss showed no meaningful improvement over five consecutive epochs.

### Validation and Metrics

For robust validation, we employed an external dataset that mirrors a realistic clinical environment, thereby assessing the model’s generalizability and robustness—essential qualities for any automated diagnostic tool. While the training cohorts were composed of research datasets with standardized acquisition protocols, clinical practice often exhibits greater variability. To bridge this gap, we utilized a dataset of T1 MPRAGE MRI scans collected from the Johns Hopkins Hydrocephalus Clinic. This dataset was initially processed using FreeSurfer’s recon-all pipeline for region-of-interest (ROI) segmentation.

To quantify model performance, we developed custom evaluation metrics specifically tailored for landmark localization. The Percentage of Correct Keypoints (landmarks) at a 3-, 5-, and 10-mm threshold (PCK@mm) is defined as the percentage of landmarks for which the Euclidean distance between the predicted and ground truth locations is less than the specified distance. In addition, we introduced PCK@V, which quantifies the percentage of Keypoints (landmarks) located within their corresponding FreeSurfer-derived ROIs. Complementing these metrics, we report the mean Euclidean distance in mm between the predicted and ground truth landmarks. These comprehensive metrics provide a rigorous evaluation of our model’s accuracy and its applicability in clinical settings.

## Results

Demographic characteristics for each cohort are summarized in Table 1. A total of 14,472 T1-weighted MRI scans from 6,299 participants were included across four datasets. The ADNI cohort provided 10,000 scans from 4,677 individuals (mean age 70.30 ± 10.6 years; 2,333 males [49.9%], 2,344 females [50.1%]) for model pretraining. For training and internal validation, we used 2,886 scans from 1,032 BLSA participants (mean age 73.32 ± 13.36 years; 471 males [45.6%], 561 females [54.4%]) and 1,391 scans from 395 BIOCARD participants (mean age 64.22 ± 11.02 years; 161 males [40.8%], 234 females [59.2%]). Finally, 195 scans from 195 Johns Hopkins Hydrocephalus Clinic patients (mean age 71.54 ± 8.30 years; 105 males [53.8%], 90 females [46.2%]) were reserved for external validation and visualization.

**Table 1.** Demographic characteristics of participants included in the training and validation datasets. Age represents participants’ age at the time of MRI acquisition demonstrated by Mean and Standard Deviation, with distributions stratified by age and gender.

| Cohort | Number of samples | Number of Participants | Age | Sex | Use |
| --- | --- | --- | --- | --- | --- |
| ADNI | 10,000 | 4,677 | 70.30 (10.6) | Male = 2,333 (49.88%)<br>Female = 2,344 (50.11%) | Pretraining |
| BLSA | 2,886 | 1,032 | 73.32 (13.36) | Male = 471 (45.63%)<br>Female = 561 (54.36%) | Training and Internal validation |
| BIOCARD | 1,391 | 395 | 64.22 (11.02) | Male = 161 (40.75%)<br>Female = 234 (59.24%) | Training and Internal validation |
| Johns Hopkins Hydrocephalus Clinic | 195 | 195 | 71.54 (8.30) | Male = 105 (53.8%)<br>Female = 90 (46.1%) | External validation and visualization |

The model was first pretrained for 50 epochs—using the same training hyperparameters and supervised learning protocol—on the ADNI dataset. We then loaded these pretrained weights and trained and validated the network on the BIOCARD and BLSA cohorts. The model reached convergence after approximately 30 training epochs. As summarized in Table 2, overall landmark localization error averaged 2.32 ± 0.41 mm in internal validation and 2.37 ± 1.52 mm in the external cohort. At a 3 mm threshold, the percentage of correct keypoints (PCK@3 mm) was 78.5% internally and 80.5% externally; increasing the threshold to 5 mm raised PCK to 96.9% and 94.4%, respectively, while PCK@10 mm exceeded 98% in both settings. When evaluating volume-based accuracy (PCK@V) in the external set, 94.8% of landmarks fell inside their anatomically defined 3D regions.

**Table 2.** Performance metrics of the proposed model on the internal and external validation datasets include Mean Euclidean Distance (MED) □ and □ Standard Deviation (SD) between predicted and annotated landmarks, Percentage of Correct Keypoints within a 10 mm threshold (PCK@10), and Percentage of Correct Keypoints within the defined 3D segmentation volume (PCK@V).

| Dataset | Internal Validation |  |  |  |  | External Validation |  |  |  |  |  |
| --- | --- | --- | --- | --- | --- | --- | --- | --- | --- | --- | --- |
| keypoint | MED | SD | PCK@3m | PCK@5m | PCK@10m | MED | SD | PCK@3m | PCK@5m | PCK@10m | PCK@V |
| overall | 2.32 | 0.41 | 78.46 | 96.92 | 99.49 | 2.37 | 1.52 | 80.51 | 94.36 | 98.46 | 94.81556 |
| Corpus Callosum Posterior | 2.7 | 2 | 86.67 | 95.38 | 97.44 | 2.21 | 2.33 | 88.72 | 95.38 | 97.95 | 93.85 |
| Corpus Callosum Anterior | 2.58 | 2.25 | 74.87 | 91.28 | 97.95 | 2.28 | 3.37 | 86.67 | 95.9 | 98.97 | 90.77 |
| Right Putamen | 2.47 | 1.44 | 72.31 | 94.87 | 100 | 2.47 | 2.65 | 81.03 | 93.85 | 97.95 | 96.41 |
| Left Putamen | 2.56 | 1.09 | 72.31 | 97.95 | 100 | 2.34 | 2.37 | 83.59 | 94.87 | 97.95 | 95.9 |
| Left Caudate | 2.06 | 1.21 | 83.08 | 98.97 | 100 | 2.17 | 2.16 | 85.13 | 96.41 | 98.46 | 97.44 |
| Right Caudate | 2.14 | 2.08 | 77.95 | 93.85 | 98.97 | 2.65 | 2.85 | 75.9 | 93.85 | 97.44 | 93.85 |
| Left Thalamus | 2.83 | 1.51 | 61.03 | 92.31 | 100 | 2.38 | 1.64 | 78.46 | 95.9 | 99.49 | 97.44 |
| Left Pallidum | 2.22 | 0.98 | 83.08 | 100 | 100 | 1.97 | 1.67 | 89.23 | 96.92 | 99.49 | 97.95 |
| Left Hippocampus | 2.83 | 1.38 | 63.59 | 93.33 | 99.49 | 2.51 | 2.32 | 74.36 | 94.36 | 98.46 | 95.9 |
| Left Amygdala | 2.43 | 0.91 | 77.44 | 99.49 | 100 | 2.29 | 2.15 | 85.13 | 95.9 | 98.97 | 97.44 |
| Right Thalamus | 1.97 | 1.06 | 87.69 | 99.49 | 100 | 2 | 1.51 | 85.64 | 97.44 | 98.97 | 98.46 |
| Right Pallidum | 2.24 | 0.96 | 82.05 | 100 | 100 | 2.23 | 1.66 | 84.1 | 96.41 | 98.97 | 96.92 |
| Right Hippocampus | 2.84 | 1.48 | 65.64 | 93.33 | 99.49 | 2.6 | 2.46 | 75.9 | 91.28 | 97.44 | 94.36 |
| Right Amygdala | 2.24 | 1.06 | 82.56 | 99.49 | 100 | 2.21 | 2.22 | 83.59 | 94.87 | 98.97 | 96.92 |
| Optic Chiasm | 1.73 | 1.01 | 94.36 | 99.49 | 100 | 1.92 | 1.82 | 90.26 | 97.44 | 98.97 | 68.21 |
| Midbrain | 1.66 | 1.01 | 94.87 | 99.49 | 100 | 1.87 | 1.68 | 90.77 | 95.9 | 98.97 | 98.97 |
| Pons | 1.83 | 1.01 | 92.31 | 99.49 | 100 | 2 | 1.94 | 91.28 | 97.44 | 98.46 | 98.97 |
| Medulla | 2 | 1.18 | 85.13 | 99.49 | 99.49 | 2.34 | 3.05 | 88.21 | 96.41 | 97.95 | 96.92 |
| Posterior Commissure | 2.07 | 1.42 | 81.03 | 95.9 | 99.49 | 2.09 | 2.75 | 86.67 | 92.31 | 95.9 | NA |
| Anterior Commissure | 2.57 | 1.22 | 67.69 | 98.46 | 100 | 2.65 | 1.82 | 68.21 | 97.44 | 99.49 | NA |
| Left Internal Capsule Genu | 2.09 | 1.19 | 80 | 98.97 | 100 | 2.3 | 1.57 | 79.49 | 96.92 | 99.49 | NA |
| Right Internal Capsule Genu | 2.18 | 1.21 | 76.92 | 98.97 | 100 | 2.62 | 1.67 | 65.64 | 96.92 | 98.97 | NA |
| Intraventricular Foramen | 2.15 | 1.13 | 80 | 98.97 | 100 | 2.44 | 1.52 | 73.33 | 97.44 | 99.49 | NA |

Region-specific analysis (Table 2) demonstrated consistently high performance across diverse structures. Posterior and anterior corpus callosum landmarks yielded mean errors of 2.70 mm and 2.58 mm internally (2.21 mm and 2.28 mm externally) with PCK@10 mm above 97% in both cohorts. Subcortical nuclei—including putamen, caudate, thalamus, and pallidum—showed internal mean errors between 2.06 mm (left caudate) and 2.83 mm (left thalamus), and external errors ranging from 2.17 mm to 2.47 mm. At 10 mm tolerance, these landmarks achieved virtually 100% PCK in internal validation and over 95% PCK externally, with volume-based accuracy exceeding 95% for all but one structure.

Limbic landmarks (hippocampus and amygdala) also performed robustly: internal errors of 2.43– 2.83 mm translated to external errors of 2.21–2.51 mm, with PCK@10 mm above 97% and PCK@V above 95%. Brainstem and optic structures were localized with exceptional precision— the midbrain and pons averaged <2 mm error and reached 100% PCK@10 mm in internal tests, and external PCK@V of 98.97%. The optic chiasm, while maintaining sub–2 mm mean error internally, showed slightly lower volume accuracy (68.2% PCK@V), reflecting its small size and variable shape.

Finally, commissural and internal-capsule landmarks exhibited mean errors of 2.07–2.65 mm internally and 2.09–2.65 mm externally, with PCK@10 mm above 95% and consistent external volume accuracy where applicable. Together, these results confirm that our model achieves sub–3 mm localization error and high robustness across anatomical sites and imaging sources.

Furthermore, we benchmarked our proposed model against published 3D landmark-detection methods including, 3D UNet(23), VNet(24), and ResUNet3D(25), using the same datasets, and the results are summarized in Table 3. As shown, our approach achieves lower mean localization error and higher PCK across multiple distance thresholds, consistently outperforming all baseline methods in multiclass landmark detection.

**Table 3.** Quantitative evaluation of BrainSignsNET versus published state-of-the-art baseline methods on the same brain MRI dataset for detecting 23 anatomical landmarks. Performance metrics include Mean Euclidean Distance (MED) and Standard Deviation (SD) between predicted and annotated landmarks, and Percentage of Correct Keypoints (PCK) across multiple Euclidean distance thresholds.

| Models | MED | SD | PCK@3mm | PCK@5mm | PCK@10mm |
| --- | --- | --- | --- | --- | --- |
| UNet (23) | 5.44 | 1.98 | 28.38 | 58.26 | 87.23 |
| VNet (24) | 4.48 | 1.44 | 36.12 | 0.69116 | 94.72 |
| ResUNet3D (25) | 3.63 | 1.25 | 49.6 | 0.80967 | 97.55 |
| **BrainSignsNET** | **2.37** | **1.52** | **80.51** | **94.36** | **98.46** |

## Discussion

In this study, we introduce BrainSignsNET, a supervised deep learning architecture specifically designed for three-dimensional landmark detection in 3D medical imaging data. We demonstrate its utility by training the model on brain T1 MPRAGE MRI scans to detect 23 anatomical landmarks, thereby establishing a robust foundation model that can be further fine-tuned to accurately localize specific anatomical structures in 3D MRI scans. The foundation model was developed and internally validated using 3D T1-weighted MPRAGE MRI scans from the ADNI, BIOCARD, and BLSA studies, with landmarks automatically extracted via FreeSurfer’s analysis as described in our methods section. External validation was subsequently performed on manually annotated scans from the Johns Hopkins Hydrocephalus Clinic, confirming the model’s generalizability across different datasets.

Our proposed model architecture is inspired by the Stacked Hourglass model introduced by Newell et al.(26), modified to effectively handle the volumetric nature of 3D data through 3D convolutional neural network (CNN) blocks. To enhance the precision of landmark localization, we integrated a soft-argmax operation, facilitating differentiable, sub-voxel accuracy during training by enabling the model to train on the exact landmark coordinates as well as the heatmaps.

Additionally, the introduction of an attention decoder branch significantly improves the model’s performance by generating a single-class attention heatmap that guides the localization process. This attention mechanism allows the model to selectively focus on regions most relevant to landmark prediction, thereby improving accuracy and robustness under challenging conditions. The resulting architecture not only achieves accurate landmark detection but also enhances interpretability through visualization of attention-guided heatmaps, aiding in model analysis and debugging.

Considering the inherent variability in anatomical structures and potential errors introduced during FreeSurfer’s analysis and landmark extraction procedures, the external validation demonstrated high accuracy in localizing anatomical landmarks, reflected by the notably low mean Euclidean distances between predicted landmarks and annotated ground truths. Precision was particularly high for well-defined single-point structures, such as the anterior commissure (AC) and intraventricular foramen. Conversely, greater localization errors were observed in anatomically complex structures, such as the corpus callosum and the caudate nucleus, likely attributable to variability in training annotations and inherent morphological diversity affecting automated annotations. To comprehensively evaluate model performance for volumetric anatomical landmarks, we introduced an additional metric quantifying the percentage of predicted landmarks located within corrected volumetric masks derived from FreeSurfer segmentation. This analysis revealed comparatively lower accuracy for small-volume or narrow structures, notably the optic chiasm and corpus callosum, while demonstrating higher accuracy in larger, structurally consistent regions like the thalamus and brainstem architectures. Importantly, these findings were validated using clinical T1-weighted MPRAGE MRI scans from elderly patients (age >70) attending the Johns Hopkins Hydrocephalus Clinic, many presenting with enlarged ventricles and brain atrophy, thereby underscoring the model’s robustness and potential applicability across diverse clinical populations.

Previous studies in medical-image landmark detection have reported mean absolute errors (MAE) between 3 and 9 mm(14–18)—substantially higher than the sub–3 mm precision achieved by our model. Moreover, these methods were typically validated on high-quality, research-acquired scans with uniform protocols, leaving their performance on noisy or pathology-laden clinical images uncertain. In contrast, our landmark-detection framework was expressly tested on an external clinical cohort from the Johns Hopkins Hydrocephalus Clinic and maintained robust accuracy (MAE ≈ 2.3 mm and PCK@3 mm ≈ 80%). Notably, an externally validated multi-class brain landmark study reported a 3 mm MAE and only 58% PCK@3 mm(19)—underscoring the precision and generalizability of our approach for brain MRI landmark detection in clinical settings. Moreover, segmentation-based approaches—such as our initial landmark extraction from FreeSurfer-derived masks—are inherently limited by the accuracy of the underlying segmentation. For landmarks without dedicated segmentation labels (e.g., the anterior commissure [AC], posterior commissure [PC], intraventricular foramen, and genu of the internal capsule), localization relies on indirect methods that introduce additional noise. In contrast, our model was trained and validated on manually refined keypoints and learns to detect these anatomical landmarks directly from image intensities and spatial context. By incorporating extensive three-dimensional augmentations during training and achieved robust performance on unseen scans acquired with diverse MRI protocols and scanner settings.

Accurate anatomical landmark detection in brain MRI scans is crucial for a variety of imaging tasks, including registration, alignment, and quantitative analyses.(1, 3, 27) Several studies have developed automated methods for measuring different indices in MRI scans. For example, one study proposed an automated technique to measure the callosal angle (CA) using segmentation-derived volumetric masks from FreeSurfer analysis; in that work, the centroid and anterior portions of the lateral ventricles were employed to align the MRI scans, with a constant pitch applied to minimize discrepancies between the automated alignment and the standard AC-PC plane alignment used in callosal angle measurements. However, these approaches can be adversely affected by anatomical changes, particularly in individuals with atrophy or hydrocephalus, as these conditions can significantly alter the shape and centroid of the lateral ventricles. Moreover, other studies using automated, atlas-based registration methods have similarly noted that brain abnormalities can impair alignment accuracy.(28–30) Our proposed method offers a validated and explainable approach to registration that can be trained on any specific landmarks—including the anterior and posterior commissures (AC and PC)—thus providing an alignment that closely adheres to standard evaluations of any brain MRI indices.

Beyond brain MRI, accurate landmark detection significantly enhances other medical imaging fields such as breast tomosynthesis (3D mammography), molecular imaging, and musculoskeletal imaging. For instance, in breast tomosynthesis, identifying anatomical landmarks like the nipple aids in lesion tracking, ensuring consistency across sequential scans and improving diagnostic precision.(31) In molecular imaging techniques like PET/CT or SPECT,(32, 33) landmark-based registration using deep brain nuclei or cardiac apex and base facilitates accurate quantitative analyses, such as tracer uptake localization in epilepsy and myocardial perfusion imaging.(34) Similarly, musculoskeletal imaging utilizes landmark detection to quantify the Cobb angle in scoliosis or automate spinal alignment in 3D CT or MRI, highlighting the broad applicability of landmark detection at various scales of clinical imaging.(35–38)

This study has several limitations. Firstly, although the datasets originated from multiple sites, they primarily represent aging and Alzheimer’s disease populations and thus lack broader representation across different age groups. Various pathological conditions, such as tumors or lesions causing structural abnormalities, were not included and may potentially reduce the generalizability and performance of our model. Evaluating the model’s performance in more diverse clinical populations remains necessary. Secondly, training was conducted exclusively on T1-weighted MPRAGE sequences, which, despite providing excellent anatomical detail, are not routinely employed for many clinical scenarios due to higher costs and limited availability. Future studies should extend this architecture’s application to more conventional MRI sequences commonly used in clinical practice.

In conclusion, our deep learning–based model achieved robust performance in detecting neuroanatomical landmarks on external-validation clinical scans, demonstrating its readiness for integration into neuroimaging pipelines for both research and clinical data. Future work could build on this foundation by incorporating multi-stage cascaded architectures or patch-based refinement strategies—using our proposed model as a backbone—to further boost detection accuracy and deliver even more precise landmark localization.

## Data Availability

The complete source code for this project is publicly available in the project's GitHub repository. Raw data used in the associated paper can be obtained upon request from the respective database.

https://github.com/SiavashShirzad/BrainSignsNET

## Data Sharing Statement

The complete codebase for BrainSignsNET, including all scripts and implementations of the 3D landmark detection network, is publicly available at https://github.com/SiavashShirzad/BrainSignsNET.git. The pretrained model weights used in this study will be provided upon reasonable request from the corresponding author and are planned to be released publicly in a future update.

## Acknowledgments

This research was supported in part by the Intramural Research Program of the National Institutes of Health (NIH). The contributions of the NIH author(s) were made as part of their official duties as NIH federal employees, are in compliance with agency policy requirements, and are considered Works of the United States Government. However, the findings and conclusions presented in this paper are those of the author(s) and do not necessarily reflect the views of the NIH or the U.S. Department of Health and Human Services.

