## Supplementary Figure 1 for "BrainSignsNET: Deep Learning-Based 3D Anatomical Landmark Detection in Human Brain Imaging"

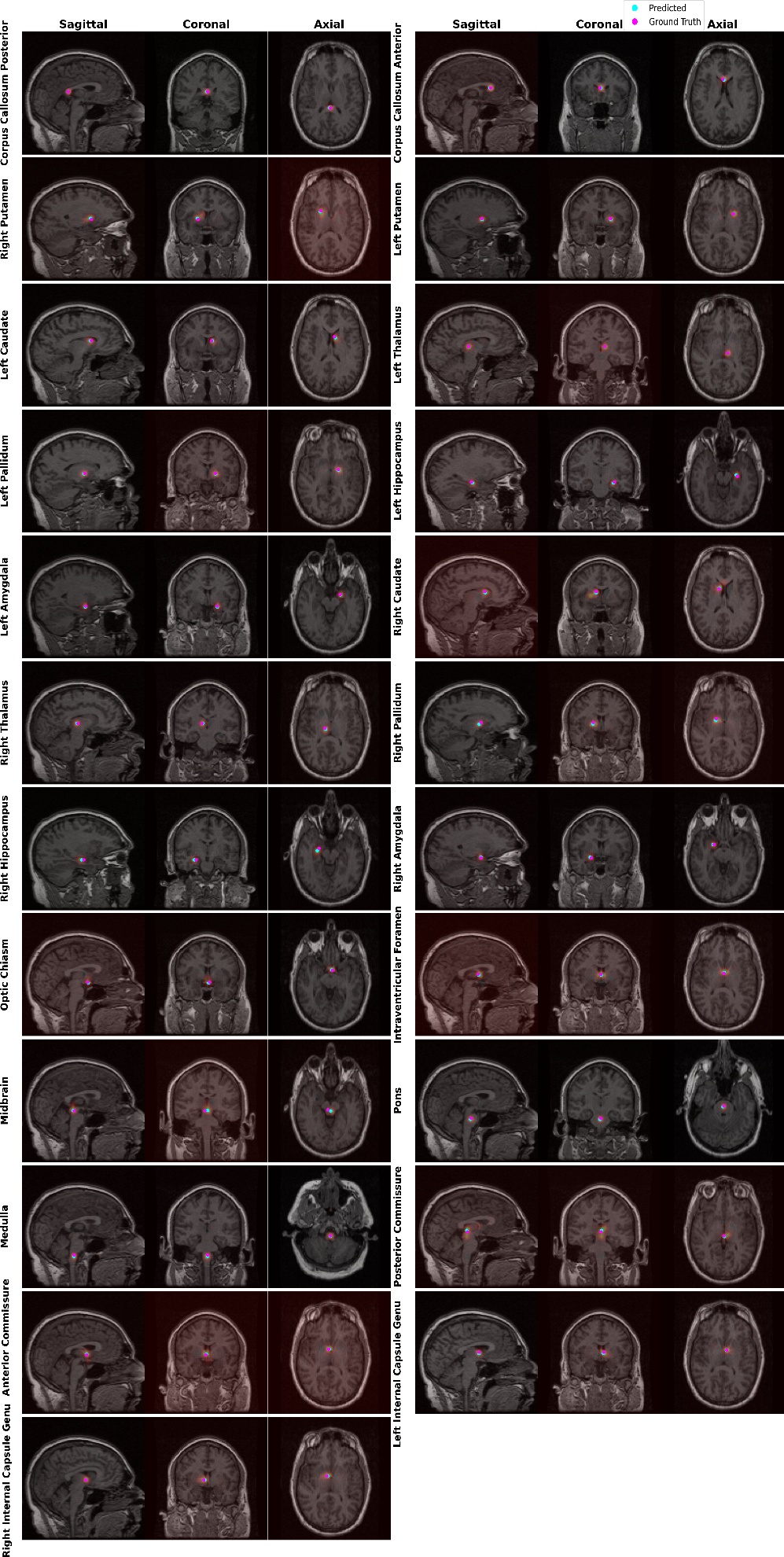

Supplementary Figure 1. Example output of the proposed model, illustrating the multiclass heatmap predictions. Predicted landmark coordinates (blue dots) and ground-truth annotated landmark (pink dots) are overlaid for visual comparison.
